# Evaluating the efficacy and cost-effectiveness of a digital, app-based intervention for depression (VMood) in community-based settings in Vietnam: A stepped-wedge cluster randomized controlled trial

**DOI:** 10.64898/2026.03.12.26348290

**Authors:** Leena W Chau, Liying Yang, Emanuel Krebs, Hui Xie, Vu Cong Nguyen, Hai Nhu Tran, Thi Thanh Xuan Nguyen, Harry Minas, Raymond W Lam, Jill K Murphy, Jeannine Ho, Kanna Hayashi, Van Nguyen Hoi, To Duc, John O’Neil

## Abstract

Depression contributes substantially to the global burden of disease, with large treatment gaps, particularly in low- and middle-income countries (LMICs) like Vietnam. Digital mental health interventions (DMHIs) offer a promising solution, yet evidence for the efficacy and cost-effectiveness of digitally delivered adaptations of evidence-based interventions in LMICs remains limited. This trial evaluated the efficacy and cost-effectiveness of VMood, an app-based DMHI for depression adapted from an evidence-based in-person intervention, in Vietnam.

We conducted a stepped-wedge cluster randomized controlled trial across 48 communes in eight Vietnamese provinces. A total of 480 adults screening positive for moderate depression (Patient Health Questionnaire-9 score 10-19), with a retention rate of 97.9%. Communes were randomized to immediate access to VMood or to enhanced usual care, consisting of a limited version of the app during the delay period. The primary outcome was depression caseness (PHQ-9 ≥ 10). Analyses followed an intention-to-treat approach using generalized estimating equations. A cost-effectiveness analysis estimated incremental costs per quality-adjusted life-year (QALY) gained.

A total of 477 trial participants were included in the primary analysis. VMood was associated with a 59% reduction in the odds of depression at 6 months (adjusted odds ratio 0.41, 95% CI 0.19–0.89). Mean PHQ-9 scores decreased by 1.9 points (95% CI −3.6 to −0.3) at 6 months. Incremental costs were 1.205 million VND ($47 USD) (95% CI: 1.006 million VND, 1.297 million VND) with 0.008 incremental QALYs (95% CI: 0.006, 0.010), resulting in a 99.7% cost-effectiveness probability at a willingness-to-pay threshold of two times GDP/capita.

VMood significantly reduced depressive symptoms and was highly likely to be cost-effective. As a scalable, low-cost intervention, VMood may help reduce the depression treatment gap in settings with limited specialist capacity, supporting investment in evidence-based DMHIs within community-based mental health systems.

This trial was registered at ClinicalTrials.gov (NCT05783531) on March 8, 2023. Available from: https://clinicaltrials.gov/study/NCT05783531.

**Author Summary:** Depression is a leading cause of disability worldwide, yet access to effective treatment remains limited, particularly in low- and middle-income countries (LMICs). Digital mental health interventions (DMHIs), such as mobile apps, can reduce depressive symptoms, especially when adapted from evidence-based treatments and delivered with human support. However, most evidence comes from high-income countries, with limited data on efficacy and cost-effectiveness in LMICs. VMood is a mobile app adapted from supported self-management, a modified approach to psychotherapy that is grounded in principles of Cognitive Behavioural Therapy and delivered in-person with support from non-specialist providers. The in-person intervention was shown to be effective in the Vietnamese context in a clinical trial. Here, we conducted a stepped-wedge cluster randomized controlled trial across eight provinces in Vietnam to evaluate the efficacy and cost-effectiveness of VMood. Participants received either the VMood app with supportive coaching provided by a social worker, or enhanced usual care, with subsequent access to the VMood app. Over six months, VMood significantly reduced depression caseness compared with the control group. Economic analysis showed that VMood represents good value for money and was feasible to deliver through existing community systems. Findings provide novel and policy-relevant evidence demonstrating that DMHIs can reduce depression and provide good value for money in LMICs.

## Introduction

Mental illnesses are a leading contributor to the burden of disease globally, accounting for significant morbidity, disability, and economic loss [1]. Depression is projected by the World Health Organization (WHO) to become the single largest disease burden worldwide by 2030 [2]. Currently, more than 300 million people are affected by major depressive or anxiety disorders, the two most prevalent mental disorders globally [3].The annual economic costs attributable globally to mental illnesses is estimated at USD $1.15 trillion [4]. Returns on investment for focusing on scaling up treatment of depression and anxiety have been suggested to be substantial, with an estimated 5.7 benefit-to-cost ratio in low- and middle-income countries (LMICs) when factoring in the value of health and economic benefits [4].

Improving equitable access to mental health care is thus a critical global priority. This is especially pertinent in LMICs like Vietnam, where care is unavailable for up to 90% of the population who need it [5]. Recent evidence shows the prevalence of mental disorders in Vietnam is 10.2% (10.1 million persons), with the prevalence of depression and anxiety being 3.0% and 2.7% (2.7 and 2.9 million persons [6]) respectively, based on the population of 98.9 million in 2021 [7]. Despite this need, less than half of all provincial health systems have at least one psychologist or psychotherapist in their public health system, and there were only three mental health nurses per 100,000 population (0.003%) as of 2020 [8]. Furthermore, most of the psychiatric workforce is concentrated in central- and provincial-level health facilities, with limited outreach to rural communities [8]. Research shows that the shortage of mental health professionals and other related mental health services will persist into at least the next decade [9]. Other persistent barriers include limited community-based care, low mental health awareness, and pervasive societal stigma that impacts help seeking [10].

Digital mental health interventions (DMHIs), including smartphone-based apps, are increasingly central to reducing the mental health treatment gap, demonstrating potential to deliver accessible, clinically effective, and scalable care across diverse populations, especially in low-resource settings [11]. Several systematic review and meta-analysis on economic evaluations of DMHIs for mental health interventions have also provided compelling evidence for their cost-effectiveness [12]. However, none of the included studies evaluated DMHIs for depression in LMICs, underscoring the substantial gap in the cost-effectiveness evidence-base for these settings.

VMood is a DMHI (mobile app) for depression that was adapted from an in-person Supported Self-Management (SSM) intervention developed in Canada. SSM is a modified approach to psychotherapy that is grounded in principles of Cognitive Behavioural Therapy and utilizes task-sharing, whereby non-specialist providers deliver psychosocial interventions. SSM is delivered through an *Antidepressant Skills Workbook (ASW)*. Results from a randomized controlled trial in community-based settings in Vietnam showed those who received SSM reduced their odds of having depression by 58% compared with the control group (care as usual) [13], with patients SSM and the associated ASW were subsequently adapted to a digital format to support accessibility and scalability, undergoing rigorous fidelity [14] and usability testing. Due to restrictions on in-person care amidst the COVID-19 pandemic and the Government of Vietnam’s fiscal changes, SSM and the associated ASW was subsequently adapted to a digital format, undergoing rigorous fidelity [14] and usability testing.

The purpose of this trial was to evaluate the efficacy and cost-effectiveness of VMood for depression in Vietnam. Specifically, we hypothesise that the VMood app is superior to an enhanced control condition (app version delivering basic health information only) for reducing depression caseness among community-based adults screening positive for depression in Vietnam, and that the DMHI will be cost-effective.

## Methods

### Study design

We conducted the trial in accordance with the Declaration of Helsinki. All procedures were approved by the Research Ethics British Columbia (BC) Board in Canada [H21-02938] and the Institutional Review Board at the Institute of Population, Health and Development (PHAD) in Vietnam [PHAD-2022/VMOOD-01]. This trial is registered at ClinicalTrials (NCT05783531). The trial was conducted from December 2023 to June 2025.

As per the study protocol [15], this study was a stepped-wedge cluster randomized trial that took place in eight Vietnamese provinces: three northern (Thai Binh, Thai Nguyen, Thanh Hoa); two central (Khanh Hoa, Da Nang); and three southern (Hau Giang, Vinh Long, Ben Tre). At the time of trial conduct, Vietnam’s administrative structure comprised 63 provincial-level units, approximately 700 districts, and over 10,000 communes. Following a major government restructuring (June 2025), governance was streamlined from a three-tier to a two-tier system, eliminating districts and reducing the number of provinces to 34 and communes to approximately 3,200. In each province, one district and six communes (municipal subdivisions) within each district were included. The trial was conducted across 48 communes. Districts and communes were purposely selected in collaboration with the Ministry of Labor, Invalids and Social Affairs (MOLISA) to ensure variation in population composition, availability of mental health resources, and economic status. In Vietnam, responsibility for mental health service provision is shared between MOLISA and the Ministry of Health (MOH). MOLISA oversees Social Protection Centres that deliver social support and rehabilitation services, while MOH manages national psychiatric hospitals and local commune health centres [16]. For confidentiality purposes, names of districts and communes have been omitted, but are available on request.

Randomization occurred at the commune level (municipal subdivisions), with communes assigned to receive either the VMood intervention immediately or one of the two delayed intervention groups (Table 1). Participants in communes assigned to the delayed intervention groups were initially provided access to the VMood app and all screening and assessment measures, but content in the app was limited to a video presenting basic information about depression. In a typical stepped-wedge design, clusters remain in the intervention arm until the study is complete; in this study, the intervention period was three months with an additional three-month follow up period. Participants in the delayed intervention groups 1 and 2 delayed groups received the full VMood intervention in the intervention period following the control period (3 months for Delay 1 group and 6 months for Delay 2). All communes in each district began the study at the same time (ensuring identical start-time distributions across the three randomization groups) owing to practical and logistical reasons, while the entry of provinces was staggered.

**Table 1.**
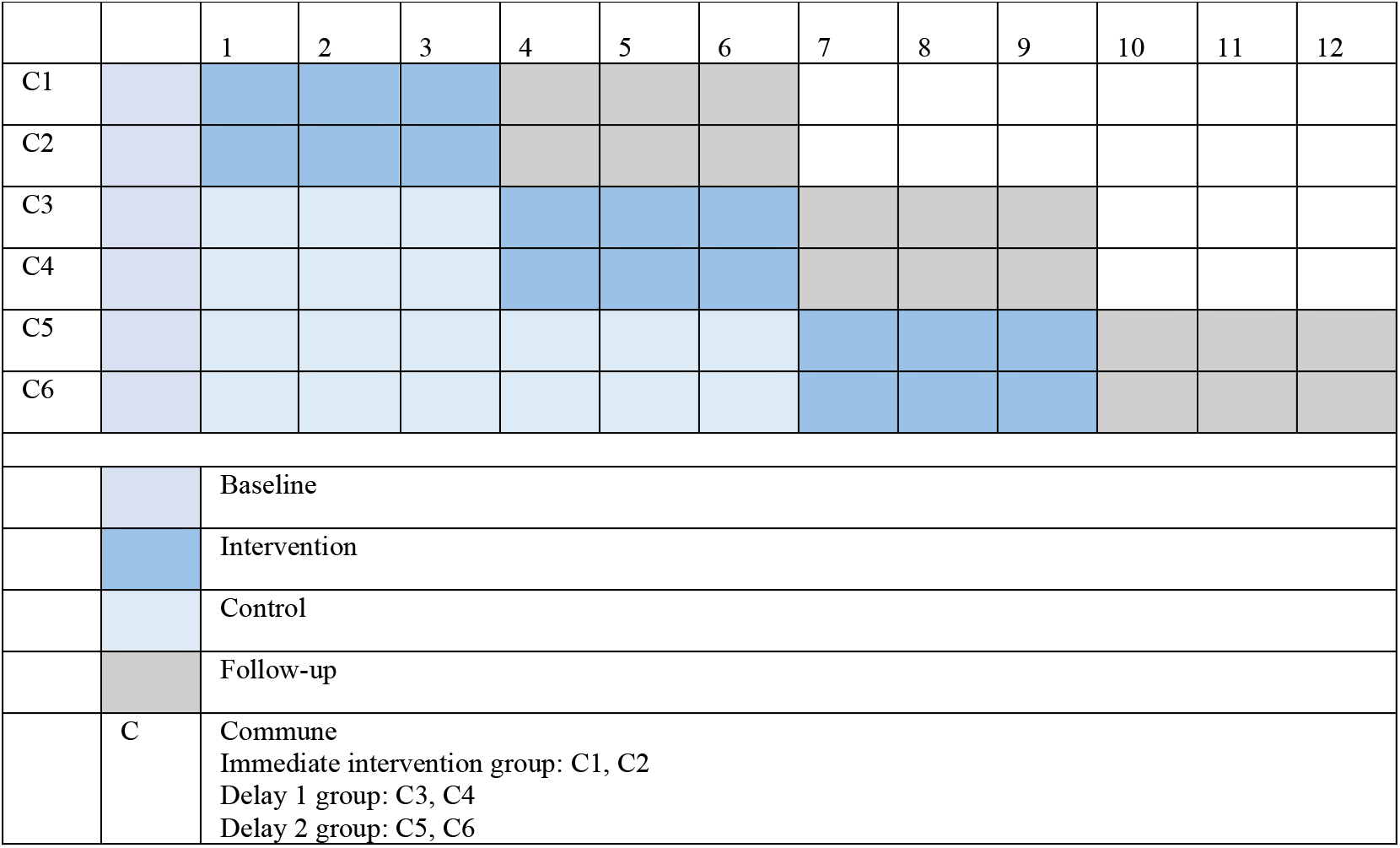
VMood stepped wedge design by month – one province/district.

We also conducted an economic evaluation alongside the trial to determine the cost-effectiveness of VMood compared to enhanced usual care. As per recommended guidelines for the conduct of CEA alongside RCTs, we obtained health resource use and health-related quality of life measures directly from trial participants and adopted an intention-to-treat design [17,18]. We adhered to best practice recommendations for the conduct and the reporting of health economic evaluations [19,20].

### Participants

Inclusion criteria included participants meeting depression caseness as determined by a PHQ-9 score of 10-19 and providing informed consent to participate in the study and complete the suite of outcomes measures at baseline, 3, 6, and 12 months. Exclusion criteria included participants experiencing severe mental illness as presented with PHQ-9 score ≥ 20, or suicidal ideation as measured by item 9 in the PHQ-9, which asks about the frequency of thoughts regarding being better off dead or hurting oneself in some way in the last two weeks. These persons were referred via an in-app notification to psychiatric care. A total of 16 individuals with severe depression were referred.

Community meetings and outreach were conducted by the study team in the communes to promote VMood. Participants entered the study in a naturalistic manner. All interested commune members were invited to download the app, complete registration, and then complete the PHQ-9. Participants completed an embedded written informed consent form via the downloaded app that enabled them to proceed to the content in the app. Participants who needed support to complete the consent form were provided contact information for the study team and were then able to contact a study team member to complete verbally by phone or by email.

### Randomization and masking

Randomization was conducted using permuted blocks to conceal allocation and was stratified by district. The randomization sequence was developed and controlled by an individual who was not involved in the study to ensure fidelity and was supervised by co-author HX. After baseline assessment, communes were randomly assigned to the three sequences in a 1:1:1 allocation ratio, stratified by district in variable block sizes using SAS v9.4 to avoid anticipated commune allocation to the treatment sequences. The outcome assessors and data analysts were blinded to group assignment. Outcome assessments were conducted by research team members not involved in the intervention delivery. Full blinding of the participants was not possible due to the nature of the VMood intervention [15].

### Procedures

VMood is comprised of 3-months’ engagement with the VMood program, which contains the core, skills-based modules from the SSM’s ASW (behavioural activation, realistic thinking, and problem solving) delivered in an interactive format using multiple modalities. Videos supplemented by text introduce each module that participants navigate through, and in-app notifications promoted engagement. For a full description of VMood and detailed procedures, refer to the protocol paper [15].

A social worker from DOLISA, the provincial-level agency of MOLISA, provided support on basic depression and app navigation remotely via the app as part of their regular job responsibilities. PHAD, a Vietnam-based non-profit research and technological organization and the team’s implementing partner, developed and delivered an online training program for social workers. PHAD also supervised development of the VMood app in collaboration with DEHA Digital Solutions, a local software and app development company.

Participants in the communes randomized to the delayed groups (two groups; Delay 1 and Delay 2) were informed via the app that they are queuing for the intervention and in the meantime received enhanced treatment as usual consisting of access to the VMood introductory modules consisting of the in-app screening measure (PHQ-9, with tracking) and the introductory video about depression. Access to the core skills-based modules was not available. During this time, any individuals identified as having severe depression (PHQ-9 score ≥ 20) or suicidal ideation based (PHQ-9 item 9 score = 1) were referred to psychiatric care. Delayed groups participants were offered access to the full VMood intervention, including the core skills-based modules from the SSM, following the three-month or six-month control period (see Table 1).

### Outcomes

The primary outcome of this study was depression caseness (PHQ-9 score ≥ 10) according to the Vietnamese version of the PHQ-9 assessed at 3- and 6-months after entering the VMood intervention group. Demographic data collected included age, sex, gender, marital status, education level, employment status and classification, ethnicity, migrant status, social insurance status, and prior hospitalization due to mental illness.

Secondary clinical outcomes included changes in PHQ-9 and GAD-7 scores from baseline to 3- and 6-months post-intervention. Other clinical outcomes and measures assessed the effect of the intervention on quality of life (WHO Quality of Life-Brief, WHOQOL-BREF); changes in use of alcohol and tobacco products (Fast Alcohol Screening Test,FAST; the first two questions on the Alcohol, Smoking and Substance Involvement Screening Test, ASSIST to assess tobacco use only). Finally, the System Usability Scale (SUS) assessed the usability of the VMood intervention.

See Table 2 for the full suite of outcome measures. Note that this paper presents results on the clinical outcomes based on the PHQ-9 and GAD-7 and the economic outcomes based on the EQ-5D-5L. Refer to protocol [15] for completeness.

**Table 2:** Outcome Measures.

| Measure | Outcome | Frequency |
| --- | --- | --- |
| *Clinical Outcomes – Depression/Anxiety* |  |  |
| *PHQ-9 | Depressive symptoms | Baseline, then monthly until 3 months post intervention |
| GAD-7 | Anxiety symptoms | Baseline, then monthly until 3 months post intervention |
| *Clinical Outcomes – Quality of Life* |  |  |
| WHOQOL-BREV | Functioning | Baseline, and at every 3 months until 3 months post-intervention |
| FAST | Alcohol use | Baseline, and at every 3 months until 3 months post-intervention |
| ASSIST – adapted | Tobacco use | Baseline, and at every 3 months until 3 months post-intervention |
| *Economic Outcomes – Cost Effectiveness Analysis* |  |  |
| EQ-5D-5L | Functioning; can use this to calculate QALYs | Baseline, and at every 3 months until 3 months post-intervention |
| HEA – adapted | Costs attributable to health resource utilization | Baseline, and at every 3 months until 3 months post-intervention |
| WPAID:D | Indirect costs attributable to productivity gains | Baseline, and at every 3 months until 3 months post-intervention |
| *Usability Outcome* |  |  |
| SUS | System (VMood) usability | At 3 months, then every 3 months until 3 months post-intervention |

\*Primary outcome was depression caseness (binary outcome) at 3 months and 6 months after initiation of the VMood intervention. Other clinical (depression) outcome was PHQ-9 change scores.

**Table 2.**
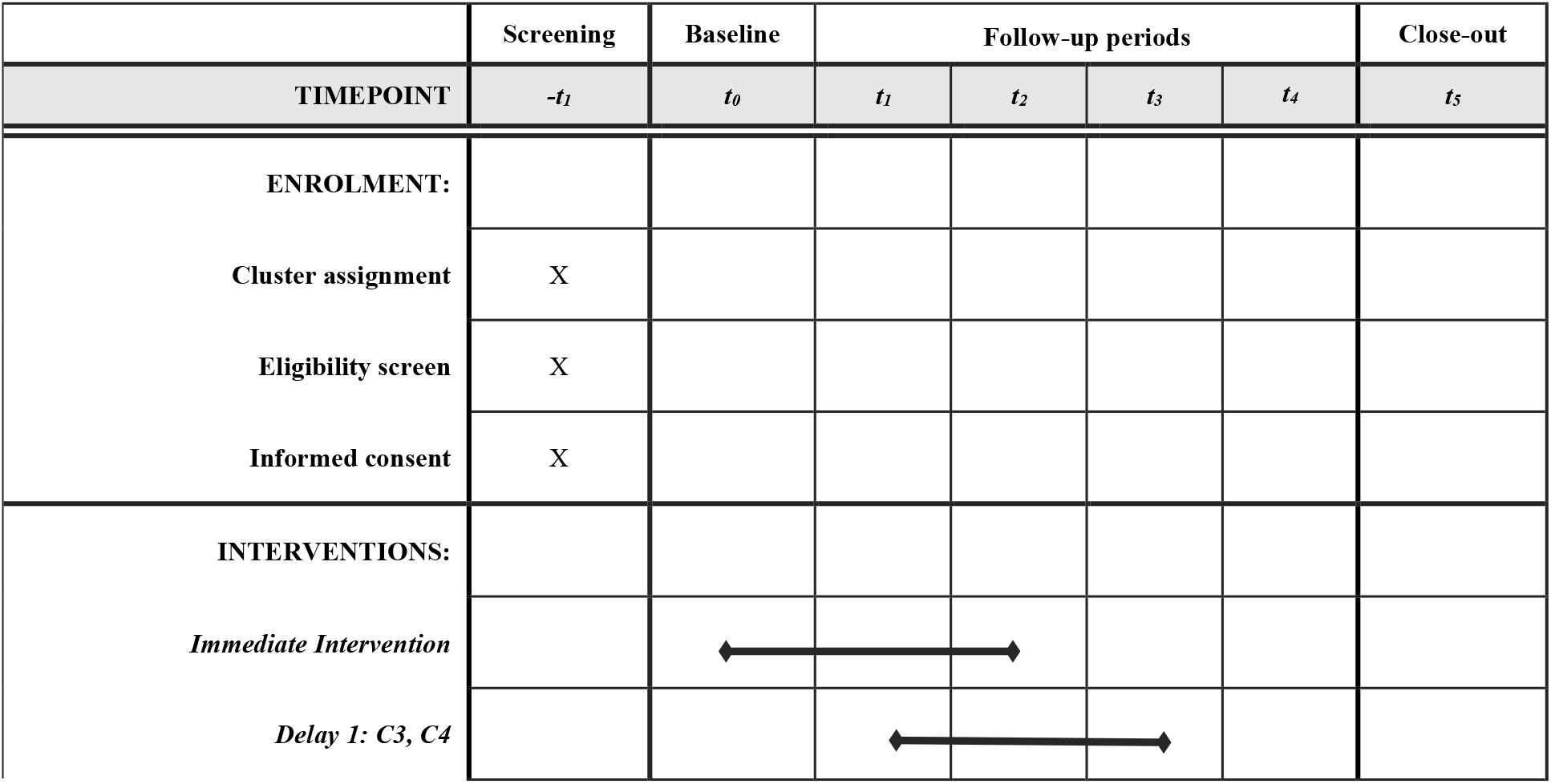
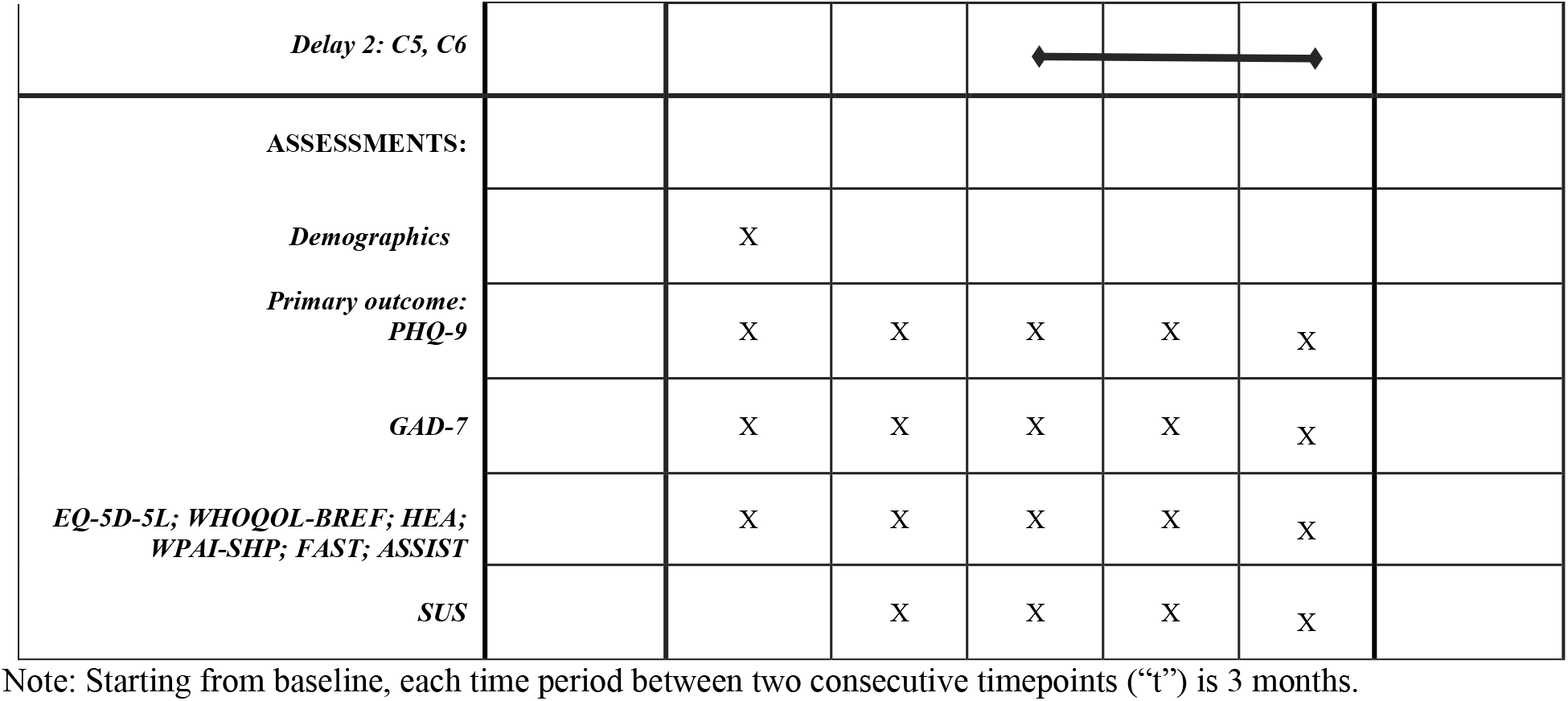
VMood SPIRIT.

Participants were administered the full survey of outcome measures at baseline (aside from the SUS), every three months until completing the intervention, and then three months after for follow-up. The full survey was administered in-person by a study team member using the REDCap online survey application [21]. See Figure 1 for the full administration schedule.

### Statistical analysis

The power calculation assumed that each commune was expected to recruit an average of eight participants and retain seven after attrition. Assuming a 42% prevalence of depression caseness (PHQ-9 scores between 10–19) in the control condition, a type I error rate of 0.05, 80% power to detect an intervention effect when the corresponding proportion in the treatment group is ≤ 30%, and an intracluster correlation coefficient of 0.01, the minimum required total sample size was 336 participants.

The primary analysis is the Intention-to-Treat (ITT) analysis that included all randomized participants. Because the assumption of Gaussian-distributed random effects was violated by the data, we employed the robust, distribution-free Generalized Estimating Equations (GEE) method [22] to estimate population-average intervention effects while accounting for the clustered, longitudinal structure of the data.

The binary primary outcome, depression caseness, measured at 3, 6, 9 and 12 months, was analysed using a logistic GEE model with the logit link. Secondary outcomes, including continuous PHQ-9 and GAD-7 scores at the same time points, were analysed using linear GEE models with the identity link. The main independent variables included indicator variables for the lengths of time since initiation of the VMood intervention to estimate intervention effects after these amounts of time post intervention initiation, a set of indicator variables for follow-up outcome assessment time points to account for secular trend, and the respective baseline outcome score. Estimates, 95% CIs, and *p* values were obtained from these GEE models. Standard errors were obtained using a commune-level bootstrap that accounts for within-cluster (commune) correlations, avoiding underestimation of standard errors in settings with small to moderate numbers of clusters and the correlation structures of participant-level longitudinal data within clusters [23].

For the binary outcome, results are expressed as adjusted odds ratios (ORs) comparing the odds of depression caseness between intervention and control periods. For continuous outcomes (PHQ-9 and GAD-7 scores), results are expressed as adjusted mean differences (Δ) between intervention and control conditions. We further computed standardized effect sizes, which were derived by dividing the log odds ratios by 1.81 for the binary outcome [24] and were calculated by dividing the adjusted mean differences by the baseline standard deviation (SD) across all participants for the continuous outcomes. All analyses were conducted using the geepack package (version 1.3.12) in R (version 4.4.1).

We conducted a range of sensitivity analyses. First, we performed analyses adjusting for baseline covariates to ensure that estimated intervention effects were not driven by residual imbalances that can arise by chance even in well-designed cluster-randomized clinical trials. We also considered an alternative model specification in which treatment exposure was treated as a continuous variable, allowing us to explore potential dose–response relationships (Supplementary Appendix S1). Second, we conducted a series of alternative analyses to assess the robustness of intervention effect estimates to protocol violations. In this trial, the amount of missing data is negligible (<2%), so the impact of data missingness is expected to be small [25]. The primary cause of protocol violations was due to participants accidentally receiving the intervention prematurely. To address this, we applied several complementary estimation approaches: (1) a per-protocol analysis, which excluded participants who violated the study protocol, thereby estimating the intervention effect among those fully adherent to the assigned protocol; (2) an as-treated analysis, which classified participants according to the treatment received rather than their randomized treatment assignment; and (3) the complier average causal effect (CACE) estimation using the two-stage residual inclusion (2SRI) approach [26]. Unlike per-protocol and as-treated analyses, the CACE analysis formally accounts for participants’ self-selection into treatment and provides a causal estimate of the intervention’s effect among individuals who would adhere to treatment assignment (Supplementary Appendices S1 and S2). Collectively, these sensitivity analyses aim to provide a thorough evaluation of the robustness and validity of the estimated intervention effects, supporting the reliability of our primary findings.

We also conducted subgroup analyses to assess treatment effects heterogeneity. For each demographic characteristic of interest, we dichotomized the variable and estimated GEE models separately for each subgroup. Differences in treatment effects across subgroups were assessed using Wald chi-squared tests of the null hypothesis that the subgroup-specific effects were equal.

### Economic analysis

For the cost-effectiveness analysis, we used data from all participants on an ITT basis with QALYs calculated using the area under the curve (AUC) methods [18,27], using a 3% annual discount rate for costs (in 2024 Vietnamese dong [VND]; societal perspective) and QALYs [19]. Direct costs attributable to health resource utilization consisted of both outpatient and inpatient services. We also derived direct costs of delivery of the intervention using existing implementation frameworks for measuring the costs of the different implementation phases for evidence-based interventions, including direct labor costs attributable to the pre-implementation (i.e., planning for delivery), implementation (i.e., delivery of the intervention) and sustainment (i.e., follow-up) of the VMood intervention [28–30].

Indirect costs consisted of costs attributable to potential productivity loss measured using the human capital approach (multiplying days and hours lost due to depression by average wages) as it remains standard practice in economic evaluation [17]. We measured indirect costs attributable to potential productivity loss using the validated Work Productivity and Activity Impairment Questionnaire: Depression (WPAI: D) [31,32]. All costs are presented in Vietnames dong (VND) and converted to US dollars at 25,480 VND for 1 USD. We provide further details on all costs in the Supplement.

For the primary economic outcome, we estimated the number of quality-adjusted life years (QALYs) accumulated during the study from participant-reported preference-based health-related quality of life [17,19] using the EuroQoL Five-Dimension Five-Level (EQ-5D-5L). The Vietnamese version of the EQ-5D-5L has been validated in Vietnam with country-specific tariffs based on social preferences obtained from a nationally representative sample [33].

## Results

A total of 48 communes across 8 provinces in Vietnam were involved in the study, with 477 participants in our ITT sample at baseline, exceeding the required sample size to allow for attrition and missed visits. In practice, only 10 participants dropped out before their six-month visits post-intervention, reflecting excellent retention supported by the social workers’ efforts. See Figure 2 for the CONSORT Flow Diagram.

**Figure 2.**
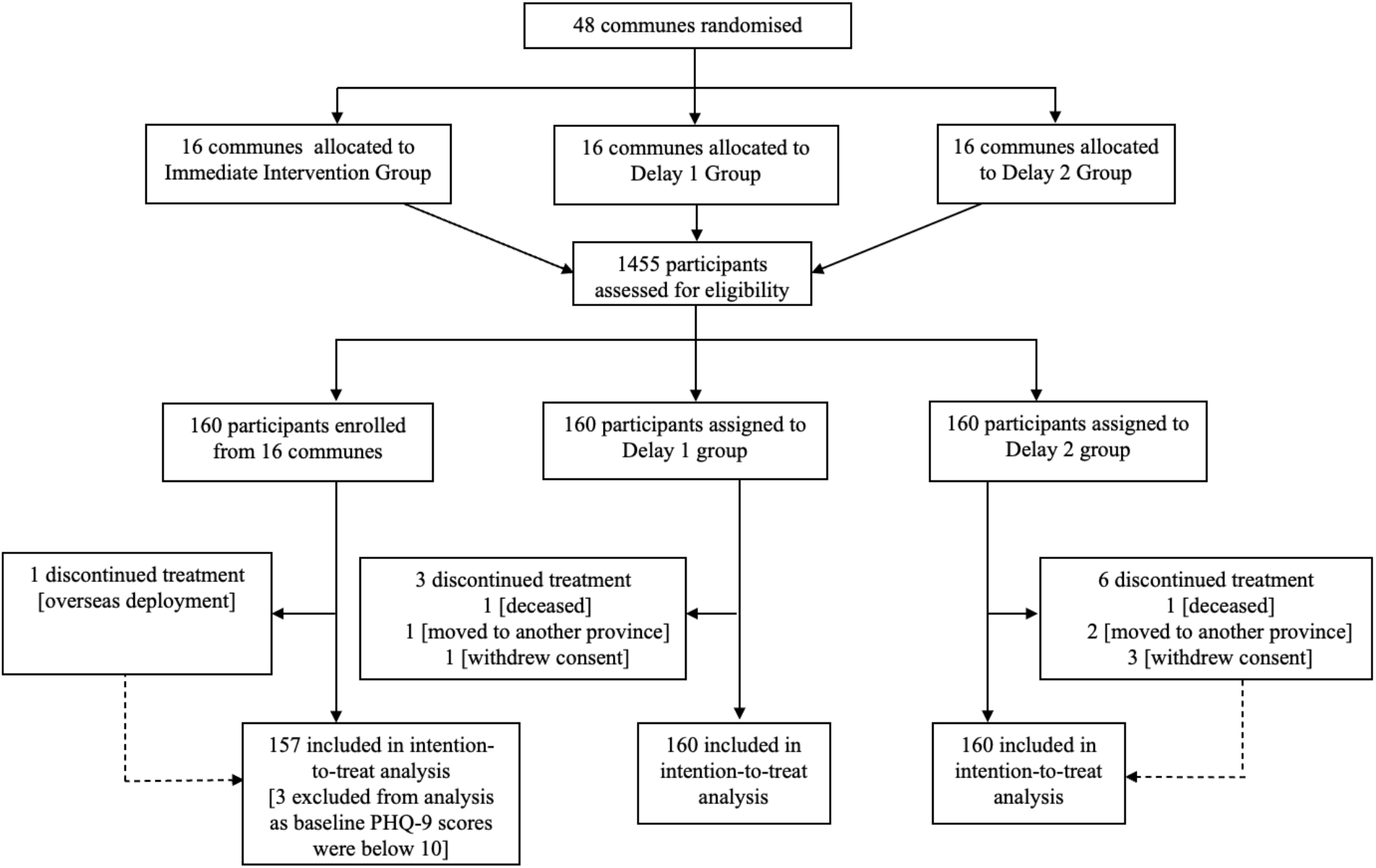
CONSORT Flow Diagram.

Due to an technical error during early software development, 118 (out of 320) participants in the delayed groups may have inadvertently gained premature access to the intervention, potentially biasing the ITT estimate toward the null. The aforementioned sensitivity analysis was therefore conducted to evaluate the reliability of our primary findings.

The baseline characteristics are summarized in Table 3. Using a chi-squared test for categorical variables, we found statistically significant differences (at the 5% level) across the three intervention groups in marital status, ethnic minority, migration status, working hours, and social health insurance coverage.

**Table 3:**
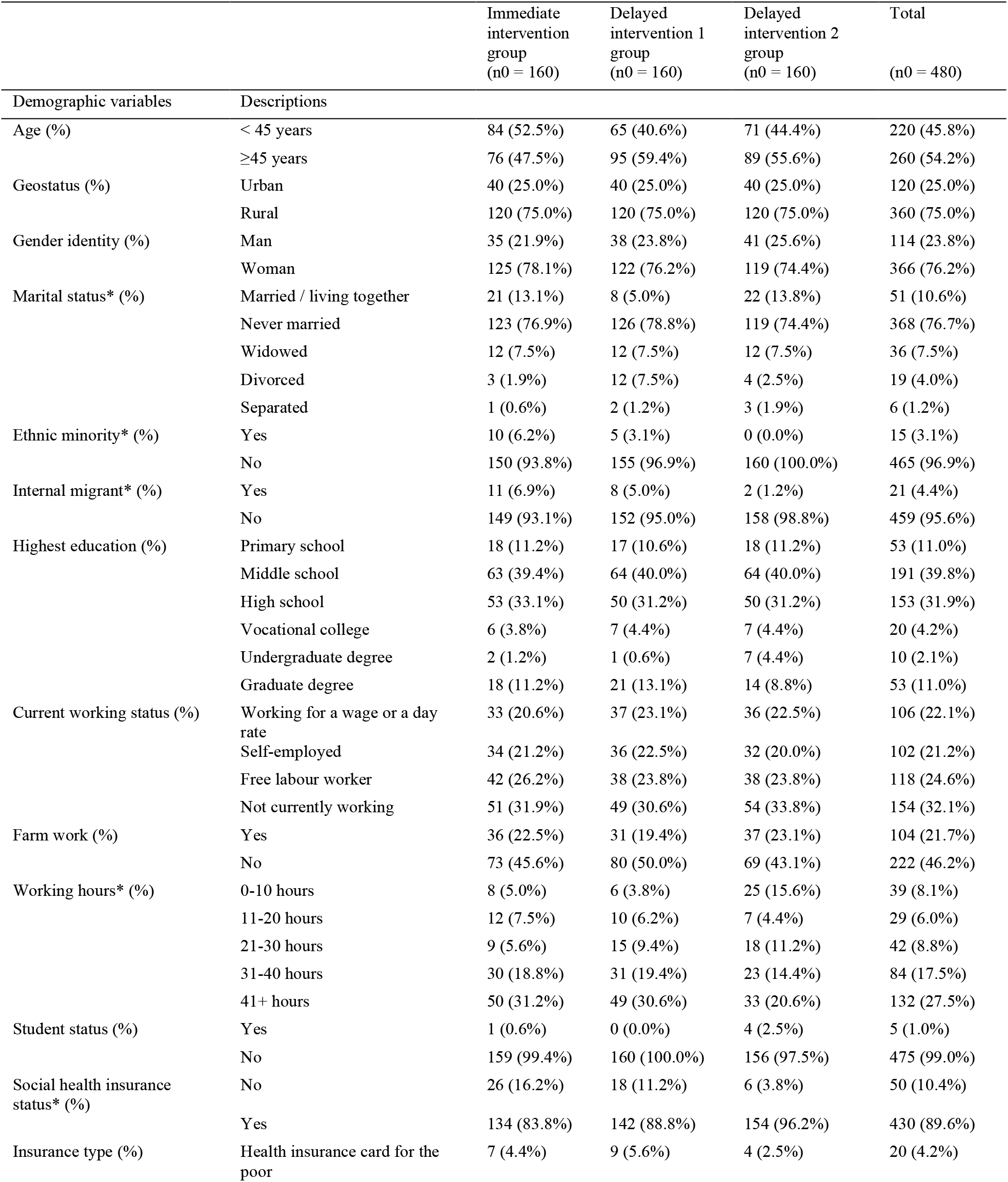
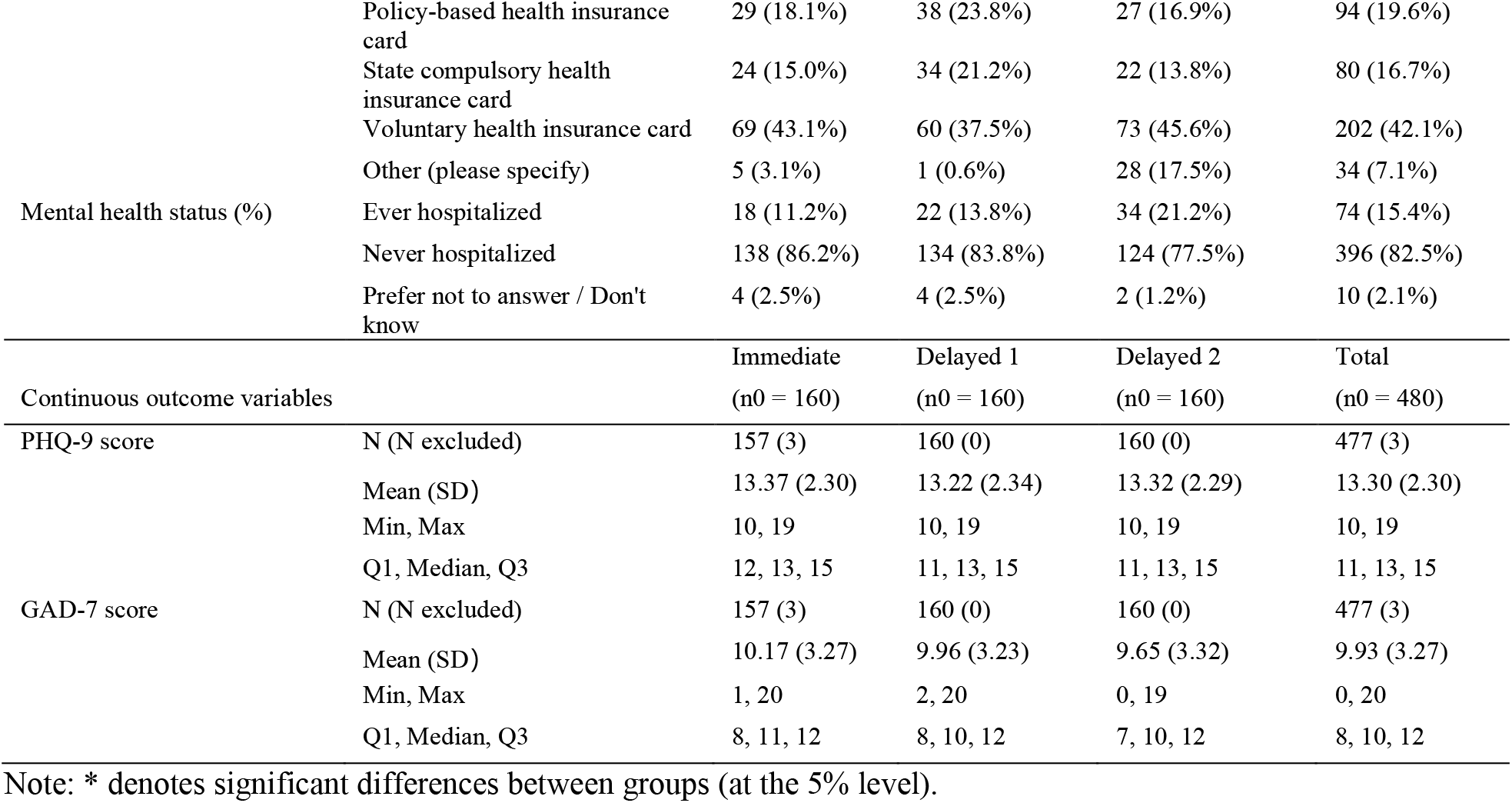
Baseline Characteristics.

### Primary analysis

The primary ITT analysis was based on data from 477 participants, after excluding three participants in the immediate intervention group whose baseline PHQ-9 scores were below 10 and therefore did not meet the inclusion criterion for moderate depressive symptoms. Including or excluding these participants did not materially change the results. During the follow-up periods, one participant from the immediate intervention group, three from the Delayed 1 group, and six from the Delayed 2 group dropped out, resulting in <2% of missingness in outcome values.

Table 4 reports the proportions of participants in the immediate intervention and delayed groups meeting depression caseness and the average PHQ-9 and GAD-7 scores across groups at different timepoints. The primary logit GEE analysis showed that participants who received the intervention had significantly lower odds of depression caseness after 6 months of the intervention than those who had not received it, whereas the estimated ITT effects at 3 months were still large but only borderline statistically significant at the 0.05 level. The adjusted odds ratio was 0.57 (95% CI 0.32-1; *p*=0.0522) at 3 months after the intervention and 0.41 (0.19-0.89; *p*=0.0217) at 6 months after the intervention, corresponding to 43% and 59% reductions in the odds of depression, respectively. For the secondary continuous outcomes, mean PHQ-9 scores decreased by 1.9 points (−3.6 to -0.3; *p*=0.0237) at 6 months, and mean GAD-7 scores declined by 2.3 points (−3.6 to -1.1; *p*=0.0003) over the same periods. The intervention thus produced substantial and sustained improvements in both depression and anxiety outcomes.

**Table 4:**
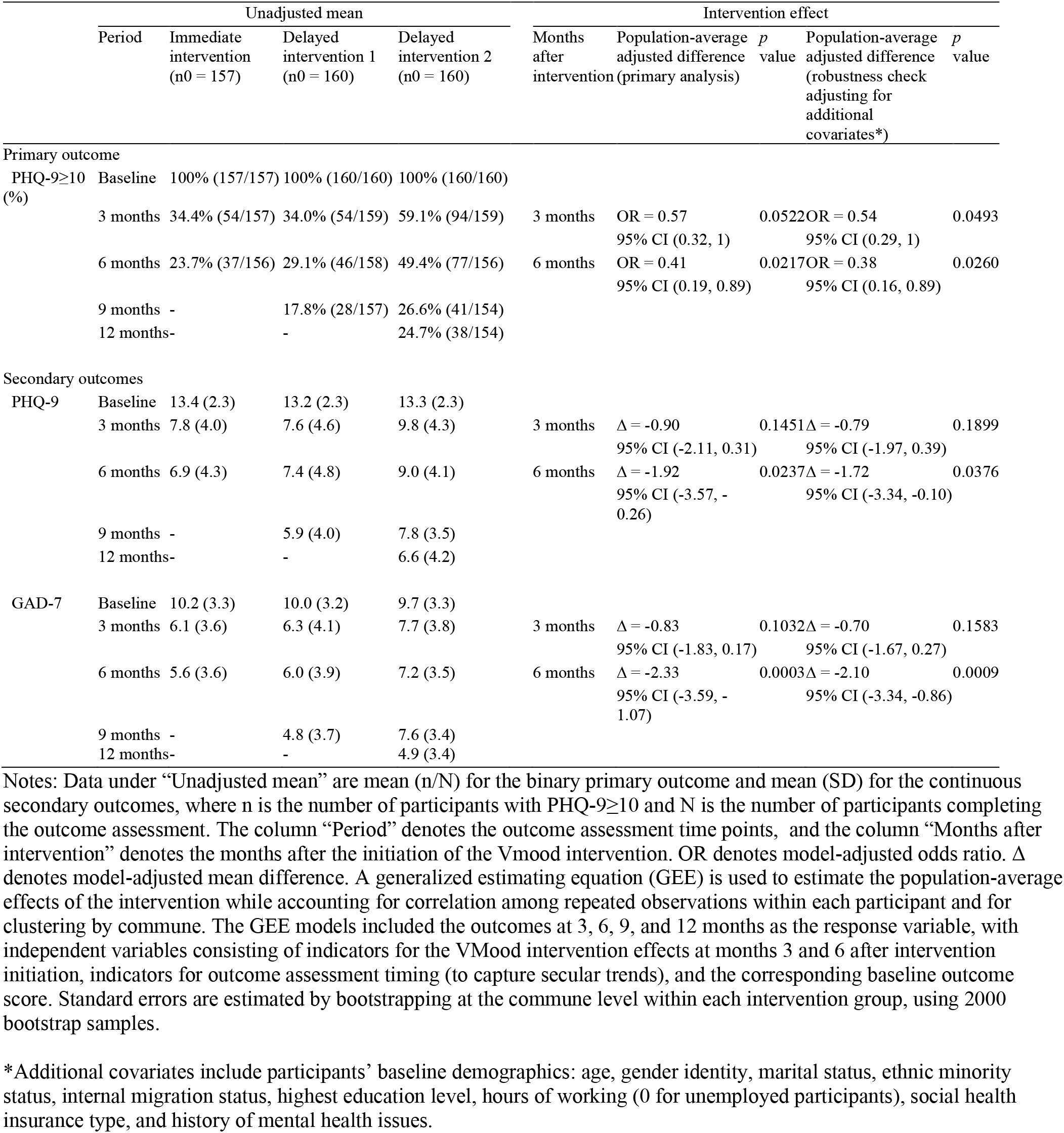
Primary Analysis: ITT Intervention Effect Estimates.

Table 5 reports the standardized effect size estimates. At 3 months, the standardized effect sizes were - 0.31 (95% CI -0.62 to 0) for depression caseness, -0.39 (−0.91 to 0.12) for PHQ-9 scores, and -0.25 (−0.56 to 0.04) for GAD-7 scores. These effects were maintained and strengthened at 6 months, reaching - 0.49 (−0.88 to -0.1), -0.83 (−1.54 to -0.13), and -0.71 (−1.1 to -0.33), respectively. By conventional benchmarks, these represent medium-to-large and clinically meaningful improvements in depression and anxiety among participants 6 months after receiving the intervention.

**Table 5:**
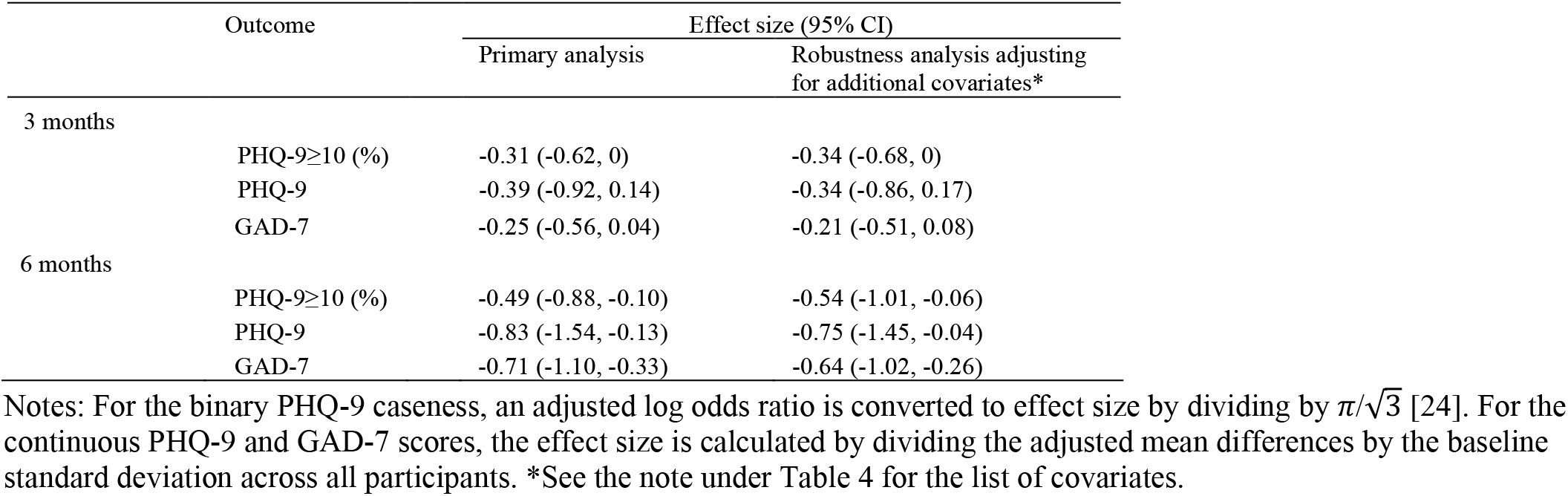
Standardized Effect Size.

### Sensitivity analysis

These findings (Tables 4 and 5) remain robust when participants’ baseline characteristics are adjusted as covariates and when treatment exposure is treated as a continuous variable to assess a dose-response relationship in GEE models (Table S1 in Supplementary Appendix S1).

For alternative estimation in the presence of protocol violations, we applied several approaches. First, excluding these individuals (per-protocol analysis) yields larger-sized estimated effects as expected, reflecting a cleaner comparison using the subsamples (Table 6). Second, classifying delayed-group participants with early access as being treated leads to attenuation of the estimated effects, likely because some may not have fully received the intervention (see Supplementary Appendix S1 for further details). Overall, for the primary outcome, the per-protocol estimate (OR = 0.36, *p* = 0.0321) represents the *strongest* estimated intervention effect—because a smaller odds ratio indicates a larger reduction in the odds of depression caseness. In contrast, the as-treated estimate (OR = 0.42, *p* = 0.0499) reflects a more *conservative* effect. Together, these estimates provide a plausible range for the 6-month average intervention impact. Moreover, using intervention assignment as the instrument and adjusting for baseline characteristics, 2SRI analyses reveal even stronger intervention effects among compliers (Tables S2 and S3 in Supplementary Appendix S1). Overall, we observe statistically significant improvements after 6 months of the intervention in the PHQ-9 caseness and in the PHQ-9 and GAD-7 scores, with large effect sizes.

**Table 6:**
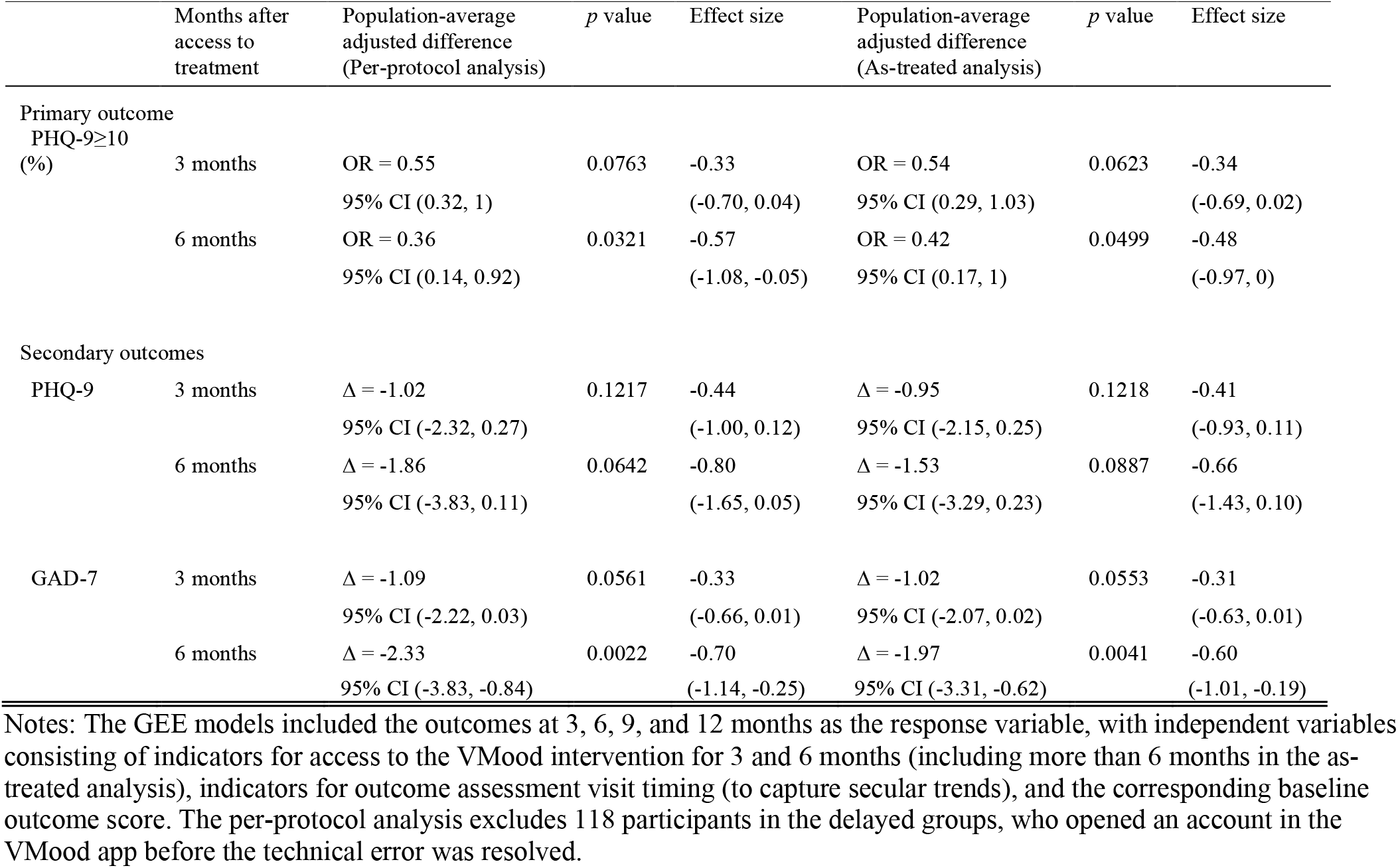
Per-protocol and As-treated Intervention Effect Estimates.

### Treatment effect heterogeneity analysis

Figure 3 shows the heterogeneity analyses of the 6-month intervention effect on the primary outcome. The estimated effects were largely homogeneous across demographic and socioeconomic groups, with no statistically significant heterogeneity by age, gender identity, educational attainment, farm work status, working hours, marital status, or health insurance coverage. The overlap in confidence intervals across these subgroups indicates that the intervention conferred comparably beneficial effects.

**Figure 3.**
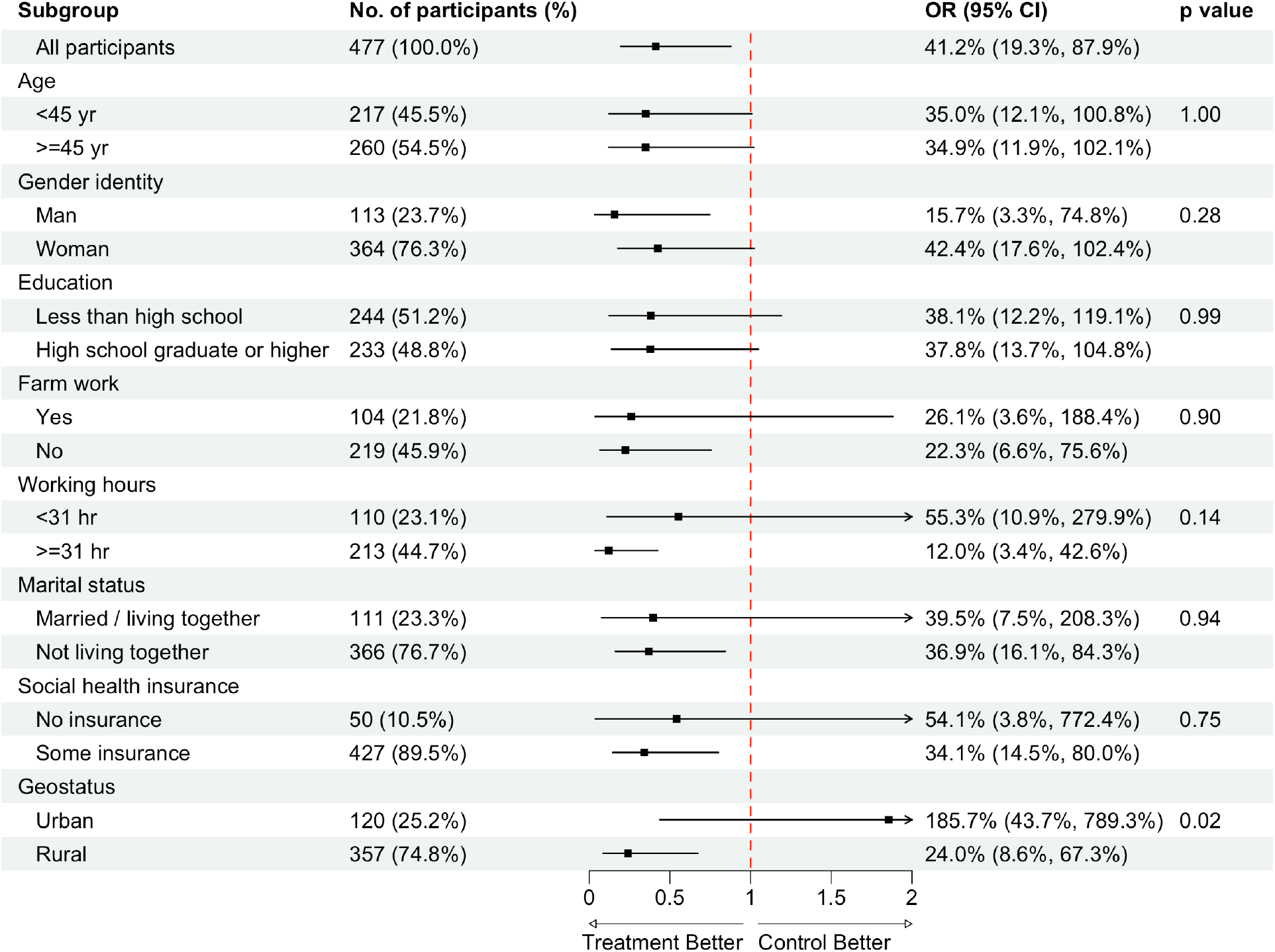
Subgroup Analysis of the 6-Month Intervention Effect on Depression Caseness.

Geostatus was the one exception (*p* = 0.02). Participants living in rural areas exhibited a markedly larger reduction in the odds of being depressed (OR = 0.24; 95% CI 0.09–0.67) compared with those living in urban areas, where the wide confidence interval, due to small sample size, indicates substantial statistical uncertainty.

For the secondary outcomes, no significant heterogeneity was detected for either the 3-month or 6-month intervention effects. Overall, these results indicate that the intervention produced consistently beneficial effects on depressive symptoms across most subgroups, with limited evidence of differential treatment effects, except for the geostatus.

### Cost-effectiveness analysis

Total mean incremental costs for VMood compared to enhanced usual care were 1.205 million VND ($47 USD) (95% CI: 1.006 million VND, 1.297 million VND; $39 US, $51 US) and mean incremental QALYs were 0.008 (95% CI: 0.006, 0.010). The ICER was estimated to be 143 million VND ($5,612 US) per QALY (95% CI: 107 million VND/QALY, 181 million VND/QALY; 95% CI: $4,199 USD/QALY, $7,104 USD/QALY), equivalent to 1.3xGDP per capita.

The INMB estimate at a willingness-to-pay threshold of 1xGDP per capita was -243 million VND (-$9,537 USD) (95% CI: -470 million VND, 78 million VND; 95% CI: -$18,446 USD, $3,061 USD), with a 2.6% probability of VMood being cost-effective (Figure 4; Panel A). The INMB estimates at a willingness-to-pay threshold of 2xGDP per capita was 719 million VND ($28,218 USD) (95% CI: 298 million VND, 1,240 million VND; 95% CI: $11,695 USD, $48,666 USD), with a 99.7% probability of VMood being cost-effective (Figure 4; Panel A).

**Figure 4.**
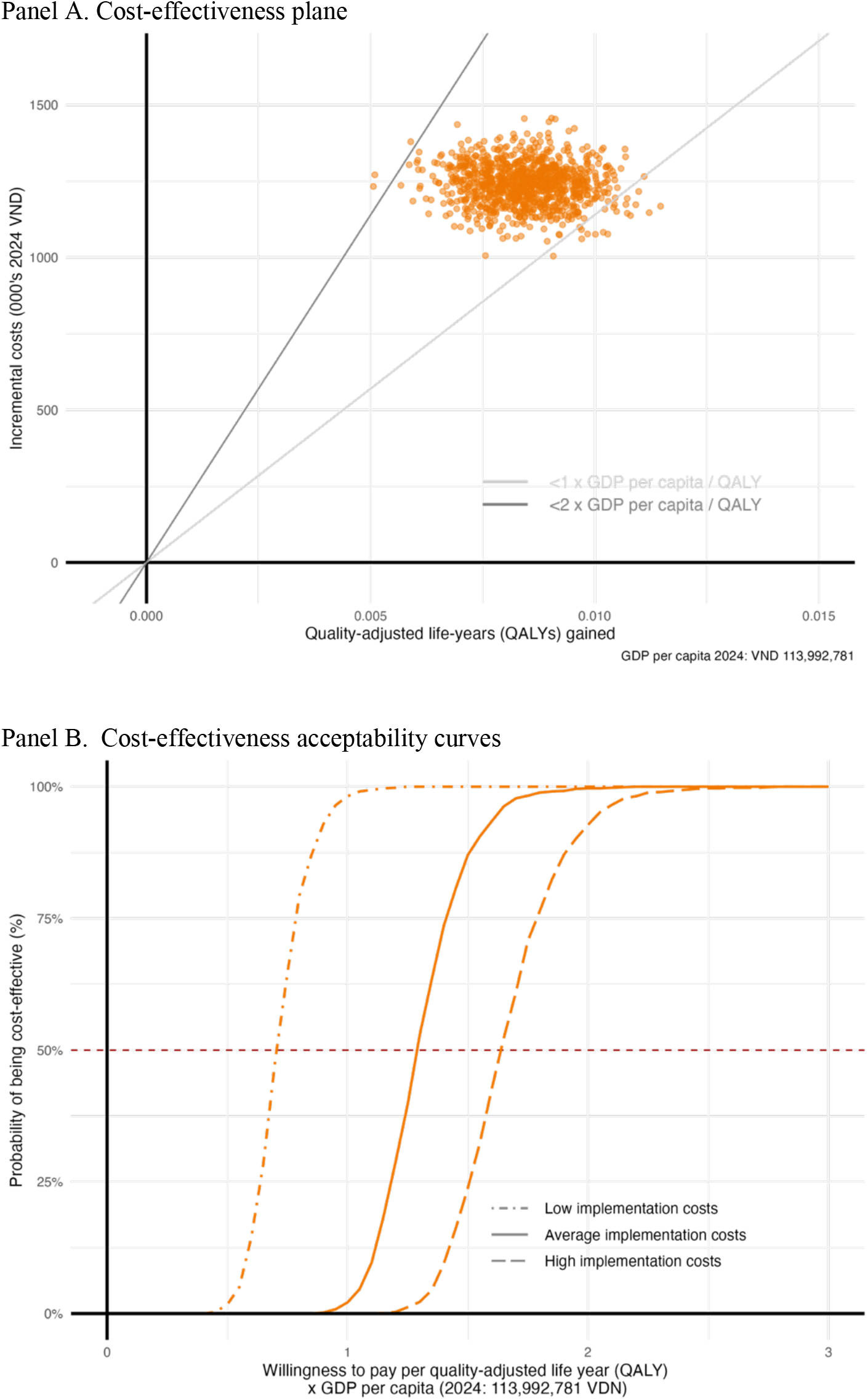
Cost-effectiveness analysis of VMood compared to enhanced usual care.

Sensitivity analysis found that with low implementation costs, there was a 98.2% probability VMood would be cost-effective at a willingness-to-pay threshold of 1xGDP per capita Figure 4; Panel B). With high implementation costs, there was a 0.0% probability VMood would be cost-effective at a willingness-to-pay threshold of 1xGDP per capita and a 92.8% probability VMood would be cost-effective at a willingness-to-pay threshold of 2xGDP per capita.

Reporting of our economic evaluation in accordance with best practice guidelines is presented in the Supplement.

## Discussion

Findings provide novel and policy-relevant evidence demonstrating that DMHIs can reduce depression and provide good value for money in LMICs. By integrating rigorous efficacy and economic evaluation within a real-world context, this study strengthens the mental health investment case for national decision makers by providing context-specific evidence on both the costs and benefits of expanding access to evidence-based, app-delivered services in resource-constrained health systems. The VMood task-sharing DMHI, rigorously adapted from an evidence-based in-person intervention shown to be effective in Vietnam [13] and with demonstrated fidelity [14], reduced depression caseness by 59% after six months and improved depression and anxiety symptom severity, while remaining cost-effective. Importantly, these findings address a critical evidence gap in LMICs, where data on scalable DMHIs and their economic value remain limited.

The consistency of the estimated intervention effects across adjusted models for different demographic and socioeconomic characteristics such as age, gender identity, and education suggests that benefits were not confined to specific demographic subgroups, supporting the potential inclusivity and scalability of the intervention. While the digital divide remains a global concern, findings suggest that DMHIs may contribute to narrowing the mental health treatment gap across different population groups, including for those who have experienced substantial barriers to accessing traditional mental health care, but this will require targeted implementation strategies to support scalability, including low-bandwidth applications that function on older smartphones, appropriate cultural and linguistic adaptation for minority communities, and providing infrastructure support.

This study fills an important gap in the evidence base on the cost-effectiveness of DMHIs in LMICs. By including country specific costs for health resources, productivity and the implementation of the VMood intervention in our economic evaluation, findings provide new evidence on the value of DMHIs that can be generalizable to other low-resource settings. We found a high probability that the VMood intervention would be cost-effective compared to enhanced usual care across a range of implementation scenarios, demonstrating that the intervention would be recommended for funding and implementation based on WHO and commonly accepted cost-effectiveness thresholds. Our findings that VMood provides good value can thus be used to inform priority setting regarding the expansion of DMHIs in LMICs.. Smartphone-based DMHIs may reduce reliance on scarce mental health professionals, lower costs associated with travel and time off work, and support task-sharing models. Training of social workers was administered via an online training program developed by PHAD, supporting future and booster training scale-up in a cost-effective manner. VMood can also be integrated into stepped-care models of care, helping to extend care to underserved populations. From a policy perspective, the demonstrated cost-effectiveness, combined with VMood’s clinical effectiveness, strengthens the case for its widespread scale-up and integration into routine settings in Vietnam.

This study has several strengths, including a clinically effective primary outcome and the integration of a health economic evaluation alongside effectiveness outcomes. The robustness of findings across sensitivity analyses adjusting for sociodemographic covariates enhances confidence in the results and supports their relevance for diverse populations. Strong cost-effectiveness results provide further support for VMood’s widespread scale-up across Vietnam and potentially in other similar resource-constrained environments, with appropriate cultural adaptation. Finally, this study is supported by a robust history of evidence demonstrating SSM’s effectiveness in Vietnam, from an initial feasibility study demonstrating the in-person SSM’s feasibility to the RCT results demonstrating its effectiveness, to this current study showing VMood’s effectiveness. Critical to the program’s success has been the long history of research-policy collaboration with the Government of Vietnam, which has facilitated policy change [34].

The study also has several limitations. While DMHIs are generally scalable, with evidence from this study indicating that the VMood program can benefit a wide range of populations, structural inequities in digital technology use persist. A recent rapid scoping review [35] showed that despite the global increase in access to smartphones and Internet, continuing challenges persist in contributing to the digital divide for certain groups, such as older people, people living in rural communities, and people living in poverty. Similarly, our recent paper found that despite widespread smartphone penetration in Vietnam, the proportion of the population owning smartphones compatible with the VMood app was limited [36]. Scale-up of VMood and other DMHIs needs to incorporate a health equity lens that focuses on reducing structural inequities and supporting more equitable access and use.

As DMHIs continue to evolve rapidly, future research should focus on emergent changes to their uptake and implementation, including around data privacy and security, policy supports, and the potential for harms in their unrestrained development and delivery. Many DMHIs are being developed that have no basis in evidence, while those adapted from evidence-based in-person formats often lack attention to fidelity across the different formats [14]. Finally, the recent accelerated growth and increasing application of artificial intelligence (AI) in various health care fields, including in mental health, may have significant implications for DMHIs for mental health [37]. While generative AI may augment DMHI delivery, concerns regarding equity, safety, and infrastructure constrain its applicability in LMICs [38]. Standalone DMHIs therefore remain highly relevant, supporting scalable and cost-effective delivery of evidence-based care using simple technologies. Future DMHI research incorporating AI should include an emphasis on diverse representation in algorithmic training data along with a detailed examination of ethical implications.

In conclusion, this study demonstrated that a task-sharing DMHI (VMood app) adapted from an evidence-based in-person intervention is clinically and cost-effective. The VMood app can serve as a compelling, low-cost and low-barrier scalable solution to address unmet mental health needs in Vietnam, and potentially other LMICs where community-based care is similarly severely limited.

## Author Contributions

Conceptualization: JON, VCN, HM, RWL, JKM, EK, LWC

Methodology: HX, EK, HM, RWL, JON, VCN, LWC

Formal analysis: LY, HX, EK

Investigation: VCN, HNT, TTXN

Resources: JON, VCN

Writing – original draft: LWC, LY, EK

Writing – review & editing: All

Supervision: JON, VCN, HM, HX

Project administration: VCN, HNT, JON, LWC, JH

Funding acquisition: JON, VCN, HM, RWL, EK, JKM, HX, KH, VNH, TD, LWC

## Acknowledgements

This research was supported by Grand Challenges Canada (R-TTS-2205-52454) and the Canadian Institutes of Health Research (PJT 178156).

## Data Availability

De-identified data will be made available following publication in the Federated Research Data Repository (FRDR). The study protocol and data dictionary will also be available. Access will be provided without restriction.

## References

1. Fan Y, Fan A, Yang Z, Fan D. Global burden of mental disorders in 204 countries and territories, 1990–2021: results from the global burden of disease study 2021. BMC Psychiatry. 2025 Dec 1;25(1):486. doi:10.1186/S12888-025-06932-Y PubMed PMID: 40375174.

2. World Health Organization. Global burden of mental disorders and the need for a comprehensive, coordinated response from health and social sectors at the country level [Internet]. Geneva, Switzerland; 2011. Report. Available from: http://apps.who.int/gb/ebwha/pdf_files/EB130/B130_R8-en.pdf

3. World Health Organization. Depression and other common mental disorders: Global health estimates. Geneva, Switzerland; 2017. Report.

4. Chisholm D, Sweeny K, Sheehan P, Rasmussen B, Smit F, Cuijpers P, et al. Scaling-up treatment of depression and anxiety: A global return on investment analysis. Lancet Psychiatry. 2016 May 1;3(5):415–24. doi:10.1016/S2215-0366(16)30024-4/ATTACHMENT/574F4366-2363-4F97-A6BA-508ABA4B68E8/MMC1.PDF PubMed PMID: 27083119.

5. Eilinghoff L, Nguyên VT, Hahn E, Nguyên VP, Lê CT, Lê TTH, et al. Changes in attitudes toward persons with mental disorders after attendance of a psychiatric curriculum among medical students in Vietnam: A cross-sectional study. Asian J Psychiatr. 2024 Mar 1;93:103949. doi:10.1016/j.ajp.2024.103949 PubMed PMID: 38335892.

6. Szücs A, van der Lubbe SCC, Arias de la Torre J, Valderas JM, Hay SI, Bisignano C, et al. The epidemiology and burden of ten mental disorders in countries of the Association of Southeast Asian Nations (ASEAN), 1990–2021: findings from the Global Burden of Disease Study 2021. Lancet Public Health. 2025 Jun 1;10(6):e480–91. doi:10.1016/S2468-2667(25)00098-2/ATTACHMENT/9B0D54F9-9B7B-46E7-8754-88677065DECA/MMC2.PDF

7. World Bank. Population, total - Viet Nam [Internet]. 2021 [cited 2025 Jun 2]. Available from: https://data.worldbank.org/indicator/SP.POP.TOTL?locations=VN

8. Le SM, Hahn E, Tran TA, Mavituna S, Minh T, Ta T. Human Resources for Mental Health Service Delivery in Viet Nam: Toward Achieving Universal Health Coverage [Internet]. 2024 [cited 2026 Jan 28]. Report. Available from: http://hdl.handle.net/10986/41623 doi:10.1596/978-1-4648-2122-6

9. Rosenberg BM, Kodish T, Cohen ZD, Gong-Guy E, Craske MG. A Novel Peer-to-Peer Coaching Program to Support Digital Mental Health: Design and Implementation. JMIR Ment Health 2022;9(1):e32430 https://mental.jmir.org/2022/1/e32430. 2022 mJan 26;9(1):e32430. doi:10.2196/32430

10. Chakrabarti S. Digital psychiatry in low-and-middle-income countries: New developments and the way forward. World J Psychiatry. 2024 Mar 3;14(3):350. doi:10.5498/WJP.V14.I3.350 PubMed PMID: 38617977.

11. Kim J, Aryee LMD, Bang H, Prajogo S, Choi YK, Hoch JS, et al. Effectiveness of Digital Mental Health Tools to Reduce Depressive and Anxiety Symptoms in Low- and Middle-Income Countries: Systematic Review and Meta-analysis. JMIR Ment Health. 2023;10. doi:10.2196/43066

12. Mitchell LM, Joshi U, Patel V, Lu C, Naslund JA. Economic evaluations of internet-based psychological interventions for anxiety disorders and depression: A systematic review. J Affect Disord. 2021 Apr 1;284:157–82. doi:10.1016/J.JAD.2021.01.092 PubMed PMID: 33601245.

13. Murphy JK, Xie H, Nguyen VC, Chau LW, Oanh PT, Nhu TK, et al. Is supported self-management for depression effective for adults in community-based settings in Vietnam?: A modified stepped-wedge cluster randomized controlled trial. Int J Ment Health Syst. 2020 Feb 12;14(1):1–17. doi:10.1186/S13033-020-00342-1/TABLES/8

14. Chau LW, Vu CN, Minas H, Hayashi K, Murphy JK, Tran HN, et al. Conceptualizing and testing fidelity-adaptation in the context of developing a digital intervention for depression from an evidence-based in-person format [under review]. BMC Health Serv Res.

15. Chau LW, Murphy JK, Nguyen VC, Xie H, Lam RW, Minas H, et al. Evaluating the effectiveness and cost-effectiveness of a digital, app-based intervention for depression (VMood) in community-based settings in Vietnam: Protocol for a stepped-wedge randomized controlled trial. PLoS One. 2023 Sep 1;18(9). doi:10.1371/JOURNAL.PONE.0290328 PubMed PMID: 37669289.

16. Minas H, Edington C, La N, Kakuma R. Mental Health in Vietnam. In: Minas H, Lewis M, editors. Mental Health in Asia and the Pacific: Historical and Cultural Perspectives [Internet]. Boston, MA: Springer US; 2017 [cited 2022 Mar 21]. p. 145–61. Available from: 10.1007/978-1-4899-7999-5_10

17. Glick HA, Doshi JA, Sonnad SS, Polsky D. Economic Evaluation in Clinical Trials. Economic Evaluation in Clinical Trials. 2014 Oct 1. doi:10.1093/MED/9780199685028.001.0001

18. Ramsey SD, Willke RJ, Glick H, Reed SD, Augustovski F, Jonsson B, et al. Cost-effectiveness analysis alongside clinical trials II-An ISPOR Good Research Practices Task Force report. Value Health. 2015 Mar 1;18(2):161–72. doi:10.1016/J.JVAL.2015.02.001 PubMed PMID: 25773551.

19. Sanders GD, Neumann PJ, Basu A, Brock DW, Feeny D, Krahn M, et al. Recommendations for Conduct, Methodological Practices, and Reporting of Cost-effectiveness Analyses: Second Panel on Cost-Effectiveness in Health and Medicine. JAMA. 2016 Sep 13;316(10):1093–103. doi:10.1001/JAMA.2016.12195 PubMed PMID: 27623463.

20. Husereau D, Drummond M, Petrou S, Carswell C, Moher D, Greenberg D, et al. Consolidated Health Economic Evaluation Reporting Standards (CHEERS)--explanation and elaboration: a report of the ISPOR Health Economic Evaluation Publication Guidelines Good Reporting Practices Task Force. Value in Health. 2013 Mar;16(2):231–50. doi:10.1016/J.JVAL.2013.02.002 PubMed PMID: 23538175.

21. REDCap. REDCap: Research Electronic Data Capture. 2022.

22. Ren Y, Hughes JP, Heagerty PJ. A simulation study of statistical approaches to data analysis in the stepped wedge design. Stat Biosci. 2020 Dec 1;12(3):399–415. doi:10.1007/S12561-019-09259-X PubMed PMID: 33747241.

23. Cheng G, Yu Z, Huang JZ. The cluster bootstrap consistency in generalized estimating equations. J Multivar Anal. 2013 Mar 1;115:33–47. doi:10.1016/J.JMVA.2012.09.003

24. Chinn S. A simple method for converting an odds ratio to effect size for use in meta-analysis. Stat Med [Internet]. 2000 [cited 2026 Jan 24];19(22):3127–31. Available from: https://pubmed.ncbi.nlm.nih.gov/11113947/

25. Xie H, Heitjan DF. Sensitivity analysis of causal inference in a clinical trial subject to crossover. Clin Trials. 2004;1(1):21–30. doi:10.1191/1740774504CN005OA PubMed PMID: 16281459.

26. Cai B, Small DS, Have TRT. Two-stage instrumental variable methods for estimating the causal odds ratio: Analysis of bias. Stat Med. 2011 Jul 10;30(15):1809–24. doi:10.1002/SIM.4241 PubMed PMID: 21495062.

27. Hunter RM, Baio G, Butt T, Morris S, Round J, Freemantle N. An educational review of the statistical issues in analysing utility data for cost-utility analysis. Pharmacoeconomics. 2015 Apr 1;33(4):355–66. doi:10.1007/S40273-014-0247-6 PubMed PMID: 25595871.

28. Kwan BM, McGinnes HL, Ory MG, Estabrooks PA, Waxmonsky JA, Glasgow RE. RE-AIM in the Real World: Use of the RE-AIM Framework for Program Planning and Evaluation in Clinical and Community Settings. Front Public Health. 2019 Nov 22;7:345. doi:10.3389/FPUBH.2019.00345/FULL PubMed PMID: 31824911.

29. Saldana L, Chamberlain P, Bradford WD, Campbell M, Landsverk J. The Cost of Implementing New Strategies (COINS): A Method for Mapping Implementation Resources Using the Stages of Implementation Completion. Child Youth Serv Rev. 2014;39:177–82. doi:10.1016/J.CHILDYOUTH.2013.10.006 PubMed PMID: 24729650.

30. Bowser DM, Henry BF, McCollister KE. Cost analysis in implementation studies of evidence-based practices for mental health and substance use disorders: a systematic review. Implement Sci. 2021 Dec 1;16(1). doi:10.1186/S13012-021-01094-3 PubMed PMID: 33706780.

31. Reilly MC, Zbrozek AS, Dukes EM. The validity and reproducibility of a work productivity and activity impairment instrument. Pharmacoeconomics. 1993;4(5):353–65. doi:10.2165/00019053-199304050-00006 PubMed PMID: 10146874.

32. Lofland JH, Pizzi L, Frick KD. A review of health-related workplace productivity loss instruments. Pharmacoeconomics. 2004;22(3):165–84. doi:10.2165/00019053-200422030-00003 PubMed PMID: 14871164.

33. Mai VQ, Sun S, Minh H Van, Luo N, Giang KB, Lindholm L, et al. An EQ-5D-5L Value Set for Vietnam. Qual Life Res. 2020 Jul 1;29(7):1923–33. doi:10.1007/S11136-020-02469-7 PubMed PMID: 32221805.

34. Murphy JK, Chau LW, Nguyen VC, Minas H, Anh DV, O’Neil J. An integrated knowledge translation (iKT) approach to advancing community-based depression care in Vietnam: lessons from an ongoing research-policy collaboration. BMC Health Serv Res. 2024 Dec 1;24(1):1–17. doi:10.1186/S12913-023-10518-3/TABLES/2 PubMed PMID: 38279141.

35. Murphy J, Khan A, Sun Q, Minas H, Hatcher S, Ng CH, et al. Needs, gaps and opportunities for standard and e-mental health care among at-risk populations in the Asia Pacific in the context of COVID-19: A rapid scoping review. Int J Equity Health. 2021 Jul 12;20(1):1–22. doi:10.1186/S12939-021-01484-5 PubMed PMID: 34253198.

36. Chau LW, Vu CN, Lam R, Minas H, Murphy JK, Tran HN, et al. Challenges and mitigation strategies in the development, usability testing, and implementation of a digital mental health intervention for depression (VMood) in Vietnam: A feasibility study. Submitted to JMIR Mental Health. 2024 Oct.

37. Lee EE, Torous J, De Choudhury M, Depp CA, Graham SA, Kim HC, et al. Artificial Intelligence for Mental Health Care: Clinical Applications, Barriers, Facilitators, and Artificial Wisdom. Biol Psychiatry Cogn Neurosci Neuroimaging. 2021 Sep 1;6(9):856–64. doi:10.1016/J.BPSC.2021.02.001 PubMed PMID: 33571718.

38. Anderson PL, Arean P, Cerezo A, Childs AW, Coppersmith DDL, et al. American Psychological Association [Internet]. 2025 [cited 2026 Jan 28]. Use of generative AI chatbots and wellness applications for mental health: An APA Health Advisory. Available from: https://www.apa.org/topics/artificial-intelligence-machine-learning/health-advisory-chatbots-wellness-apps

